# Evaluating later or expanded premises hours for alcohol in the night-time economy (ELEPHANT): statistical analysis plan

**DOI:** 10.1101/2024.05.07.24306991

**Authors:** Nurnabi Sheikh, Ines Henriques-Cadby, Houra Haghpanahan, Colin Angus, Rachel O’Donnell, Carol Emslie, Niamh Fitzgerald, Jim Lewsey

**Affiliations:** Health Economics and Health Technology Assessment (HEHTA), School of Health and Wellbeing, University of Glasgow, United Kingdom; Department of Mathematics, The University of Manchester, United Kingdom; School of Medicine and Population Health, University of Sheffield, United Kingdom; Institute for Social Marketing & Health, University of Stirling, United Kingdom; Research Centre for Health, School of Health and Life Sciences, Glasgow Caledonian University, Glasgow, Scotland, United Kingdom

## Abstract

This document provides the statistical analysis plan (SAP) for the ELEPHANT study (Evaluating later or expanded premises hours for alcohol in the night-time economy), an interrupted time series study following a natural experiment design, for evaluating the impact of late-night trading hours of alcohol premises in the night-time economy.

## 1. Background

Alcohol consumption is recognised as a substantial threat to public health and one of the underlying contributors for over 200 communicable and noncommunicable diseases [1–4]. It is a leading causal factor of preventable fatalities and is strongly linked with criminal activities, acts of violence, road traffic accidents, and various forms of injuries [2–5]. Addressing these occurrences imposes significant financial burdens on society and necessitates the allocation of substantial resources from frontline services, including ambulance, healthcare, and law enforcement [1–7].

In 2016, harmful use of alcohol led to the premature deaths of three million people globally, accounting for 5.3% of all deaths, including 0.9 million injury-related deaths attributable to road injuries, self-harm, and interpersonal violence [1]. In 2022, 1,276 people in Scotland died from an alcohol specific cause (that is, wholly attributable to alcohol) with a 2% increase from 1,245 in 2021; that is an average of 25 people every week [8]. In 2022/23, 31,206 alcohol-specific hospital admissions were recorded in Scotland; among them 28,800 were admitted in general acute units that is equivalent to 532 hospital stays per 100,000 population (age-standardised) and is about 3.4 times greater than the number of alcohol-related hospital stays recorded in 1981/82 [9,10]. A recent study estimated that 16% of ambulance call-outs in Scotland were associated with alcohol-related incidents [11], costing the National Health Service (NHS) £31.5 million in 2019 [11,12]. In 2019/2020, in Scotland, 44% of violent crimes involved offenders who were under the influence of alcohol; with 20% of victims reported drinking alcohol immediately prior to the incidents [13]. The annual cost of crimes in Scotland is estimated at >£1.5bn despite progress made in tackling violence [14].

Alcohol-related harms are prevalent in the late hours of the night (i.e., 12am to 5am), particularly on the weekends, when intoxication, fatigue, and irritability contribute to an increase in alcohol-related injuries and violence [15,16]. In northeast England, up to 70% of emergency department patients on Friday and Saturday were reported to have consumed alcohol [17]. On weekends in Scotland in 2019, the highest concentration of alcohol-related ambulance call-outs occurred between 9pm and 1am, whereas the highest concentration of non-alcohol-related call-outs appeared between 6pm and 10pm [11].

Alcohol control interventions focusing on the physical and temporal availability of alcohol are crucial and one of the most cost-effective ‘Best Buy’ interventions recommended by the World Health Organisation to reduce alcohol-related harms [18]. These interventions include enforcing strict legal age limits for purchasing and consuming alcohol, implementing licensing systems for alcohol premises, setting limits on the number of premises in an area, and restricting the hours and days of alcohol sales [7,19,20]. However, the Scottish alcohol licencing system is not set up to reduce alcohol availability; in theory, but not in practice, local licensing authorities could cap outlet numbers.

Systematic reviews find that extensions in late night opening of alcohol premises are linked to increased intoxication, crimes, injuries, and burden on public services [21–26]. A 1-hour extension of late-night trading hours in two night-life areas in the central district of Amsterdam was associated with 34% more alcohol-related ambulance call-outs and injuries (from 2am to 6am) [27]; in a study across 18 Norwegian cities, each additional 1-hour extension to opening times was associated with a 16% increase in crimes (10pm to 5am) and the converse was true for each 1 hour reduction in opening hours [28]. Alcohol-related hospital admissions among adolescents and young adults were substantially reduced in Germany and Switzerland by limiting the hours of opening of off-premise outlets (where alcohol could be purchased for off-site consumption, e.g., supermarkets, shops) [29,30]. In Newcastle Australia, late night trading hours were reduced from 5am to 3:30am with a 1:30am ‘one-way door’ (more commonly called ‘lockout’ in Australia). The reduced trading hours (not the lockout) were associated with a 74% reduction in police recorded assaults after five years [31].

The 2003 Licensing Act, for England and Wales, allowed for 24-hour licensing, when this was agreed locally for both on- and off-licence premises at the same time. The Licensing Act 2003 granted licensing authorities greater control over licensed premises, providing local communities with a more significant role in licensing decisions. Prior to the enactment of the Act, alcohol licences were issued in Magistrates’ courts. The Act introduced a shift towards flexible licensing hours, allowing premises to determine their own operating hours based on customer demand. In contrast, before the Act, venues were bound by specific opening and closing times. The extension of licensing hours was intended to foster a more European-style ‘cafe culture,’ and there was an opportunity for all those affected by the licence to make their comments on it. Early studies on 2003 Act had mixed findings in terms of Accident and Emergency (A&E) attendances and crimes but used before and after study designs and so were methodologically weaker than the international studies [32]. Using a more robust time series approach, Humphreys and colleagues found no evidence of an overall increase in violent crime in the City of Manchester but found a statistically significant 36% increase in violence between 3am and 6am [32]. This suggested that violence shifted to later time periods. However, they did not consider wider health outcomes and pointed to the need for future research “*to move beyond black-box evaluative designs by investigating the impact of policy on exposure (e*.*g*., *alcohol availability) as well as the impact of exposure on multiple indicators of physical and social harm*” [32]. A later study found that the Licencing Act 2003 had little influence on drinking occasions and alcohol harms, possibly due to previous liberalisation and greater availability of late-night drinking options [33].

In Scotland, the 2005 Licensing Act (‘the Act’) designates local authority ‘Licensing Boards’ as the decision-making body for licensing matters, such as who can sell alcohol, where it can be sold, conditions of sale, and the hours and days of sale. The Act limits the hours that off-sales venues can be open in Scotland to between 10am and 10pm. Under the 2005 Act, there is a presumption against granting 24-hour licences for on-trade premises, including for one-off events, unless there are exceptional circumstances (defined in guidance as local or national festivals held occasionally). The Act created a role in the licensing system for local public health departments based within the National Health Service. Along with the police and others, local public health departments are ‘statutory consultees’ who are informed of all applications for new or amended premises licences (including changes to opening hours). Any changes to local policy on trading hours are reflected in ‘Statements of Licensing Policy’, which undergo a process of public consultation. However, to our knowledge, no previous study in the United Kingdom (UK) has considered the impact of later trading hours on alcohol-related ambulance call-outs, and there have been no studies of later trading hours in Scotland for decades. The latest permitted opening hours for on-premises changed from 10pm to 11pm on weekdays in Scotland in the 1970s and were found (in a before/after survey) to have slightly increased consumption in younger women, and slightly shifted drinking to later times [34]. As outlined below, local Licensing Boards in Glasgow and Aberdeen have in recent years given permission, upon application, to premises in certain licence categories, to close later at night.

The ELEPHANT (Evaluating Later or Expanded Premises Hours for Alcohol in the Night-Time economy) study was designed to provide important and robust contextualised qualitative and quantitative evidence of the introduction, operation, and impact of these later trading hours in the two cities. While much of the existing literature has tended to concentrate on specific dimensions of alcohol-related harms, our study stands out by adopting a relatively holistic approach broadly in line with the statutory licensing objectives: preventing crime and disorder, securing public safety, preventing public nuisance, protecting children and young persons from harm, and protecting and improving public health. We aim to provide a more nuanced understanding of the broader impact of late-night trading hours of alcohol premises. The purpose of this statistical analysis plan (SAP) for ELEPHANT is to provide a comprehensive outline and description of the methods and procedure to quantitatively assess the impact of late-night trading hours of alcohol premises on the volume and geographic and temporal patterning of alcohol-related ambulance call-outs, total ambulance call-outs, and reported crimes that are plausibly alcohol-related.

## 2. Methods

### 2.1 The policy changes: later opening hours in Glasgow and Aberdeen

In Glasgow, as of 12th April 2019, under a scheme planned by the Glasgow Licensing Board (GLB), 10 nightclub premises (among 1,352 on-premises outlets) were granted a variation in their licence, enabling them to open for an extra hour from 3am until 4am. This extension was initially granted for 12 months. The scheme was described by the Licensing Board as an ‘opportunity to reward and continue to encourage great practice in the nightclub trade’ since the extra hour was only granted to premises meeting certain criteria [35]. The licence holder had to demonstrate, to the satisfaction of the GLB, not only that the premises made a positive contribution to the late-night economy, but that it invested in safety and security measures for both staff and customers and to promote the licensing objectives. The scheme was subsequently extended for the same 10 premises beyond the initial 12 months to open for an extra hour from 3am until 4am and made a permanent arrangement under the 2024 GLB local licensing policy.

In Aberdeen, changes took place from January 2017 such that, initially, premises that provide “significant entertainment”, and later any premises (both on-premises and off-premises), were allowed to request to stay open later as a result of changes in the new statement of licencing policy. From November 2018, and continuing throughout 2021, bars and pubs (which previously had to close no later than 1am) began to apply to extend their hours - some to 2am and some to 3am. A total of 86 applications approved to extend premises hours between 2017 and 2021 by the Aberdeen Licensing Board. Among them 6 premises applied multiple times and 42 premises applied for extended hours that did not include late-night (post mid-night) hours including shops, cafes, and restaurants. The remaining 38 premises with extended late-night closing hours in Aberdeen were included in this study.

### 2.2 Study design

The legislative changes regarding later opening hours of premises that sell alcohol in Glasgow and Aberdeen are considered a ‘natural experiment’ that can be evaluated. This quantitative evaluation of this natural experiment will use routine data sources to evaluate the impact of these local alcohol licensing policy changes on ambulance call-outs and reported crimes in Glasgow and Aberdeen. We will compare these outcomes before and after the implementation of the policy changes using a time series study design, considering the gradual uptake of the later hours in Aberdeen. Multiple outcomes will be evaluated and compared to assess the impact of the policy changes. We will also test for differential impact of the policy changes within sex, age, and socio-economic deprivation sub-groups. Finally, we will use Bayesian spatio-temporal models to assess whether the extended opening hours led to significant changes in the timing and/or location of outcomes.

#### 2.2.1 Interrupted time series

An interrupted time series (ITS) design will be used to evaluate the impact of the later opening hours across the period 2015 to 2022. Interrupted time series (ITS) design is a widely used method for strengthening before-after study design [36]. The implementation periods (considered from the date at which applications were approved to extend premises hours) for the later opening hours will be regarded an interruption in the time series for the ELEPHANT study, and data will be collected before and after that interruption. In Glasgow, 10 premises were granted to implement extra hour from the same date and so the date of the interruption was 12 April 2019 for Glasgow. However, the ‘interruption’ in Aberdeen occurred gradually following the approval date for the extended hours of the 38 premises. We therefore generated a cumulative exposure measure, detailed in the section on measurement of exposure variable. ITS is a non-randomised design and one of the most effective research designs to establish causality when randomized controlled trials (RCTs) are not practical [37,38]. It facilitates statistical modelling to be carried out to examine seasonality, autocorrelation, random fluctuations, and heteroskedasticity, all of which might lead to bias into an assessment of the impacts of the interruption/intervention [36,39]. In addition, we will utilise location-based control groups [40]. Control groups which not been exposed to the intervention under study have the potential to improve internal validity [41].

#### 2.2.2 Control selection

The objective of the control in an ITS study is to minimise biases caused by time-varying confounders, seasonality, or other events that occurred during the implementation of alcohol policies [38]. In this study, we will use a location-based control if an appropriate control location can be identified. In a location-based control, different geographical areas with similar features are typically chosen as a control, where the policy or intervention has not been adopted. We consider council areas in Scotland as potential control areas. The process of appropriate control selection and corresponding study design has been outlined in a flow diagram (see **Figure 1**).

**Figure 1:**
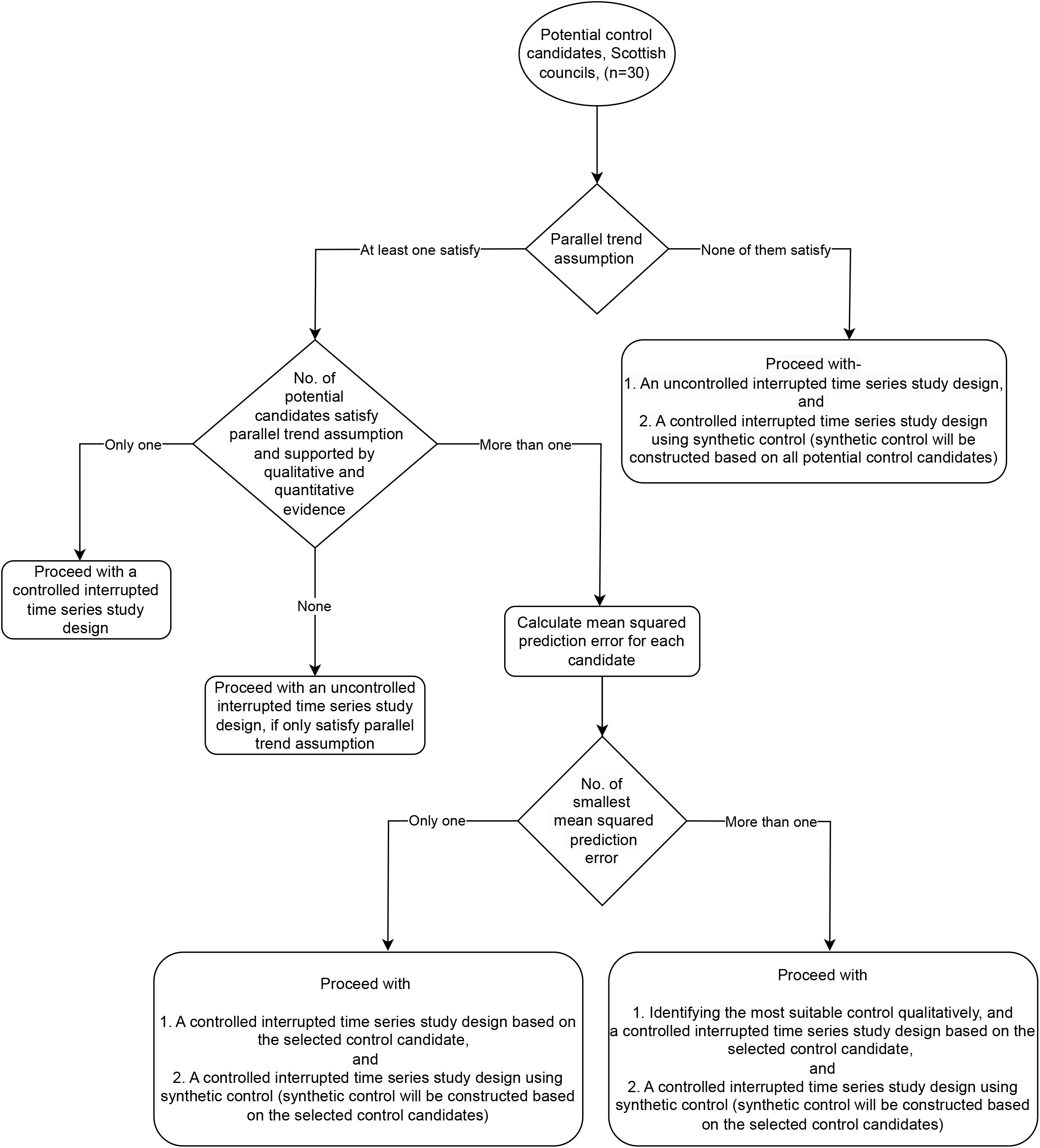
Flow diagram for the control selection procedure.

In this study, we will assess the appropriateness of the 30 Scottish councils/cities (except intervention cities-Aberdeen, and Glasgow), since no policy changes in these cities’ late night trading hours that were comparable to those in Aberdeen and Glasgow took place during our initial assessment. These cities’ late-night trading hours were temporarily changed for holidays like Christmas and New Year’s, but we expect that the intervention cities would be similarly impacted by these occasions. The appropriateness of the potential control candidates (n=30) will be tested by visually and statistically assessing the assumption of parallel trends. Graphically, we will observe the pre-intervention differences between the intervention and control groups. If the parallel test assumption holds, then the pre-intervention differences will be constant over time. There are no formal statistical tests for this assumption, and passing the parallel test does not necessarily mean that the parallel trend is true [42]. However, failing the parallel tests will make the parallel trends assumption less plausible. We will estimate the slope of the differences between the intervention and control groups in the pre-intervention periods and check whether the slope coefficient is statistically significantly close to zero. If the parallel trends assumption holds, then the slope coefficient of the differences will be significantly close to zero.

If none of the potential control candidates satisfy parallel trends assumption, we will proceed with an uncontrolled ITS study design (**Figure 1**). In addition, we will construct a synthetic control based on all potential control candidates since synthetic control does not need to satisfy parallel trends and will proceed with a controlled ITS study design based on synthetic control [43]. The synthetic control method (SCM) is widely recommended for ITS designs where multiple prospective control groups are available [38,44]. The SCM is a data-driven process that builds a control series by calculating a weighted average of potential controls [44]. To generate weights for the SCM, we will collect and utilise data from all Scottish regions outwith Glasgow and Aberdeen. The weights will be generated based on pre-intervention outcomes of potential control groups so that pre-intervention trend of synthetic control matches well with intervention (the process will be followed twice to generate synthetic control for Aberdeen and Glasgow). Along with pre-intervention outcomes, other covariates representing time-varying confounding such as per capita gross disposable household income [45], number of alcohol premises/population-adjusted alcohol premises [46], and weather (mean temperature and rainfall from the Centre for Environmental Data Analysis archive) will be used to generate weights for synthetic control. In order to determine more accurate weights for the synthetic control series, we will incorporate population size-adjusted alcohol-related ambulance call-out and reported crimes rates for all regions in Scotland. We anticipate that by utilising population-adjusted rates, the SCM series will yield a closer match to the observed outcomes prior to the intervention.

On the other hand, if at least one candidate satisfies parallel trends assumption, we will assess the qualitative and quantitative evidence on control group suitability. For example, we will qualitatively assess whether other policy changes connected with alcohol premises hours occurred at the selected control locations after our initial assessment, which might limit their suitability. The quantitative team will first conduct a qualitative assessment, and then the entire team will provide their feedback on the assessment. In addition, we will consider on-premises outlet density per 1000 population, socio-economic, and geographical features to assess control groups. If potential controls only satisfy parallel trends assumption but did not get support from the qualitative and quantitative assessment, we will proceed with an uncontrolled ITS study design. However, if only one candidate satisfies both parallel trends assumption and is supported by further evidence, we will proceed with a controlled ITS study design. We will calculate mean squared prediction errors (MSPE) for each of them, if more than one candidate satisfies both parallel trends assumption and has favourable qualitative and quantitative evidence support. If we get only one smallest MSPE, we will use that corresponding control and proceed with a controlled ITS study design. In addition, we will construct a synthetic control based on the selected control groups and will proceed with a controlled ITS study design based on synthetic control. On the other hand, if there is more than one similar smallest MSPEs (i.e., two or more very close MSPE), we will select the most suitable control qualitatively and will procced with a controlled ITS study design based on the corresponding control and will also construct synthetic control by incorporating selected control candidates and proceed only with a controlled ITS study design.

We will use a difference-in-difference approach, such that differences between intervention and control will be used as outcome variables for both primary and secondary outcomes. In order to validate our findings, we will conduct falsification tests involving time series for only the control group fitted with Aberdeen/Glasgow policy intervention points to observe any changes after the policy periods in the control group. There are more specifics about the different modelling approaches in **Table 1**.

**Table 1.** Type of modelling approaches.

| Model | Model type | Time series | Policy exposure/<br>covariate | Type of control |
| --- | --- | --- | --- | --- |
| Model 1 | Uncontrolled | Aberdeen | Aberdeen<br>(staggered) | None |
| Model 2 |  | Glasgow | Glasgow (binary) | None |
| Model 3 | Controlled,<br>difference in<br>difference (if any<br>suitable control<br>identified) | Differences between<br>Aberdeen and control<br>city | Aberdeen<br>(staggered) | Location-based<br>control |
| Model 4 |  | Differences between<br>Glasgow and control<br>city | Glasgow (binary) | Location-based<br>control |
| Model 5 | Controlled,<br>difference in<br>difference (based<br>on synthetic<br>control) | Synthetic Aberdeen (if<br>applicable) | Aberdeen<br>(staggered) | Synthetic control |
| Model 6 |  | Synthetic Glasgow (if<br>applicable) | Glasgow (binary) | Synthetic control |

### 2.3 Data source and information governance

Scotland-wide data will be obtained from two separate routinely recorded sources: ambulance call-out data from the Scottish Ambulance Service (SAS) and reported crimes data from Police Scotland. Electronic records of all ambulance call-outs in Scotland (∼500,000 per year) are available from SAS. Data includes incident number, date and time of incident, patient postcode, age-group, sex, council name, and X/Y location coordinates of incident. We will use an algorithm (which we developed in collaboration with SAS in earlier work) to optimise identification of alcohol-related call-outs using free text entries in call-out records [11]. We will also obtain anonymised individual reported crimes level data from Police Scotland IT systems for all cities in Scotland. We will utilize the Scottish Government Justice Directorate (SGJD) crime codes to identify reported crimes that are plausibly alcohol-related. Subsequently, we will gather data from the Police Scotland IT systems pertaining to these designated crime codes. Available variables include type of crimes, council name, date and time of crime and X/Y coordinates of crime location. We have made agreements for sharing information with both the SAS and Police Scotland, allowing us to retain and analyse data on secure University servers. Information for qualitative assessment will include analysis of local licensing board policy statements for 2018-2023 [47].

### 2.4 Outcome measures

#### 2.4.1 Primary outcome

The primary outcome for the ELEPHANT study is the weekly or 4-weekly totals of weekend night-time (defined as Friday 20:00 to 23:59, Saturday 00:00 to 05:59 and 20:00 to 23:59, and Sunday 00:00 to 05.59) alcohol-related ambulance call-outs [48]. We will consider weekly or 4-weekly totals of weekend night-time events, providing 52 ‘weekly’ or 13 ‘monthly’ observations per year as the unit of analysis. The decision on which level of aggregation to use for the analysis is based on consideration whether weekly units lead to problems of small/zero counts.

#### 2.4.2 Secondary outcomes

The secondary outcomes are weekly or 4-weekly totals of all weekend night-time ambulance call-outs (i.e., irrespective of attribution to alcohol or otherwise) and weekly or 4-weekly totals of weekend night-time reported crimes (using the same time cut-offs as in 2.4.1 above). The following types of recorded crime will be analysed in our study, serious assault; robbery and assault with intent to rob; threats and extortion; sexual assault; trespass, crimes against public order; drugs; disorderly content; litter offences; drunkenness; Offences by licensed persons; other offences against liquor licensing laws; and drunk driving.

## 3. Data analysis

### 3.1 Measurement of exposure variable

We will generate exposure variables separately for Aberdeen and Glasgow. In Glasgow all 10 nightclubs were permitted to implement extended hours from the same date, i.e., 12^th^ April 2019. This will therefore be reflected in our analyses using a dummy variable taking the value 0 for all time periods before 12^th^ April 2019 and 1 in all analyses thereafter.

In Aberdeen, not all venues applied for extended hours immediately, with the increase in permitted later opening hours being staggered over several years. We reflect this in our analyses by generating an exposure variable that takes values between 0 and 1 for all time periods, with 0 reflecting no extended hours and 1 reflecting the maximum permitted, possible, additional person hours. This variable therefore takes the value 0 for all time periods before 21^st^ March 2017. For later periods, we will calculate the total number of additional person hours available across all premises in a week and take this as a proportion of the maximum number of additional person hours – 127,962, which is the sum of the maximum additional person hours available per week across all 38 premises. The maximum potential additional person hours for a given venue in a week is calculated by using the formula (additional hours x the number of nights additional hours are permitted to be used in a week x capacity of the venue). For instance, if five premises with capacities of 400, 300, 350, 400, and 260 were permitted to open for two additional hours on both Friday and Saturday in a given week, the maximum potential additional person hours available for that week would be (1600+1200+1400+1600+1040 = 6840). We would then divide this by 127,962 to give an exposure measure of 0.0535 for that week.

### 3.2 Confounding variables

In interrupted time series designs, time-varying confounding is a threat to obtaining a causal estimate of an intervention effect [49]. One example of potential time-varying confounding is any other interventions identified across the time period (e.g., permitted a significant number of on-premises outlets, minimum unit pricing policy, etc.). Other potential confounders for our evaluation are per capita gross disposable household income, and weather (mean temperature, and rainfall) and we will adjust for this using quarterly gross disposable household income and weather indicators data for each local authority in Scotland [50]. Finally, we will adjust for seasonality by testing the model fit of different approaches (e.g., Fourier terms or dummy variables representing the time point of the year each weekly or 4-weekly period represents).

### 3.3 Power calculation

We have obtained a preliminary dataset of ambulance call-outs for May 2016 – April 2018, which confirms the stability and quality of the data. We estimate a pre-intervention mean of 1623 alcohol-related call-outs per 4-weekly intervals, assuming one-third of call-outs at these times are alcohol-related, and 74 and 236 in Aberdeen and Glasgow, respectively. If we conservatively assume an effect size of 6% and a Type I error probability of 5%, and autocorrelation (lag 1) of 0.2, we estimate 80% power with 36, 4-weekly pre- and 30, 4-weekly post-intervention data points [51]. If, as expected, the effect size is greater than 6% (recall that the estimates from the literature range from 16-34%) then power will be greater than 80%.

### 3.4 Statistical modelling

Initially, data will be analysed descriptively and trends along with summary statistics will be generated and displayed in tables and graphs. Following this we will carry out statistical inference as outlined below. We will construct separate time-series models for primary and secondary outcomes.

#### 3.4.1 Autoregressive integrated moving average models

We will fit autoregressive integrated moving average (ARIMA) models with the weekly or 4-weekly total counts as the outcome variables. Such models account for autocorrelation and underlying temporal trends, and we will use Akaike information criterion (AIC) and Bayesian information criterion (BIC) to determine the best fitting ARIMA models. After a best fitting model is determined, we will then add the intervention variables to measure the effect size. The following are the possible base modelling approaches, summarised in **Table 1**. We will also construct models following the sensitivity analysis, described in section 3.6.

Models 1-6 will be fitted to both alcohol-related ambulance call-outs (primary outcome), and all secondary outcomes. Models 3 and 4 will utilise a location-based control and will be based on difference in difference technique for generating counterfactuals, while models 5 and 6 will adopt a synthetic control technique. In accordance with Bernal’s recommendation, differences between the intervention and control outcomes (i.e., Aberdeen/Glasgow minus (-) control city) will be modelled if any suitable control identified [38]. In order to undertake hypothesis testing on the synthetic control method, non-parametric permutation test will be performed, and 95% confidence intervals will be constructed based on the permutation test’s p-values [52,53].

### 3.5 Subgroup Analyses

In line with previous literature [25,32], we will also fit models for different strata based on what time of day the outcomes occurred to test for effect modification by time of day (e.g., a larger increase in outcome observed between 4am and 6am when premises have closed). The differential impact by age, socio-economic status and sex of patient will be assessed by repeating the primary analysis of ARIMA models after stratifying the alcohol-related ambulance call-out data, separately, by age (< 25, 25-34, 35-44, and 45+), sex (female, male, and others), and socio-economic deprivation based on the Scottish Index of Multiple Deprivation (SIMD) scores on a scale of 1 to 10, where 1 is regarded as most deprived areas and 10 is least deprived areas. Subgroup analyses will be conducted for the primary outcome i.e., for the weekly/ 4-weekly alcohol-related ambulance callouts.

### 3.6 Sensitivity Analyses

A range of pre-specified sensitivity analyses will be conducted. If we deviate from our a priori plans, we will explain the rationale for doing so in our reporting:

- Type of intervention covariate: the definition above assumes a linear association between exposure to additional hours and outcome. We will test the appropriateness of this assumption by categorisation of the covariate and fitting of restricted cubic splines if appropriate [54].
- Type of counterfactual: test sensitivity of findings if use location-based control (control city if any and SCM) [38].
- Population size-adjusted rates: To assess consistency of our findings, we will generate population size-adjusted alcohol-related ambulance call-outs and crimes and then re-run all models.
- Number of premises exposed in Aberdeen: we will use the point in time when the policy has implemented at least half ‘strength’ (i.e., when the value of the covariate is > 0.5) as the point of policy implementation, with a dummy variable taking the value 0 for all time periods prior to this date and 1 for all periods thereafter. We will then re-run the associated models with this alternative exposure measure.
- Dummy policy covariate for Aberdeen: Separately, we will conduct a before and after analysis of the adoption of the new policy around later opening hours by using a binary variable for the Aberdeen covariate that takes the value 0 before 21st March 2017 and 1 for all subsequent periods.
- Level of aggregation: we will test the sensitivity of our results if we change the unit of analysis, for example to weekly, fortnightly, or 12-weekly counts or rates (rates will be generated by dividing ambulance calls/crimes by population at risk; population living in a city will be considered population at risk). A potential benefit of 12-weekly counts is reducing the number of zero (so that we could generate rates) or small counts when conducting the subgroup analyses.
- Boundary of cities: we will test the sensitivity of our results if we narrow the boundaries of the cities. We will expect any intervention effect to be increased if we only include areas in close proximity to where the intervention occurred.
- Length of pre-trend series: we will test the sensitivity of our results if we bring forward the start of our time series to test how well the underlying trend is being modelled.

### 3.7 Patterning of harms over time

Assessing changes in spatio-temporal patterns of harms will enable us to establish how the distribution of harms relates to the distribution of venues and the extent to which the different policy changes have changed this distribution in each city. Results from this analysis, taken alongside the results of the time series analysis will illustrate whether extending opening hours is associated with a change in the spatial distribution of harms over and above any change in their overall level.

The spatio-temporal distribution of alcohol premises, ambulance call outs and reported-crimes will be assessed by modelling the number of call outs and reported-crimes, in each area and each time period using a Poisson regression with a log link [55]. The geographic units of analysis (e.g. data zones/intermediate zones) and the time periods (e.g., monthly/quarterly/yearly) will depend on the number of incidents in each unit and period so as to ensure sufficient data is available at those levels, for each outcome. In addition, Bayesian space-time models [56–59] will be fitted to model alcohol-related ambulance call outs and reported-crimes committed in the study period. These models will include a variable reflecting the number of additional opening hours for premises within each area at each time point. These models will enable assessment of any significant spatial, temporal, or spatio-temporal (interaction) effects that explain variation in call-outs such as longer-term trends and/or intervention effects. We will further explore the existence of more complex effects and interactions, such as non-linear temporal effects and interactions between neighbouring geographical units, by fitting BYM (Besag, York & Mollie) [60] and Conditional Autoregressive (CAR) models [61]. Goodness of fit will be assessed using the Deviance Information Criterion [62].

This analysis will allow us to label geographical units (e.g. Datazones) within the study areas of Glasgow City and Aberdeen City as having increasing, decreasing or stable rates of ambulance callouts and/or reported crimes. We will also be able to identify hotspots and cold spots for alcohol-related harms and assess whether these have changed following the policy change. Additional point process analysis at point level, rather than aggregated at areal level, may be conducted, time permitting, to more naturally examine the spatial patterns without the restriction of administrative areas [64].

## 4. Software

Analyses will be conducted using the statistical software packages Stata, R, Stan, and Winbugs.

## 5. Reporting

We will present the results of our study in accordance with standard reporting guidelines for reporting observational studies [63] and previously published reports of natural experiments similar to the one we conducted. We will present time series in descriptive plots similar to the earlier literature by illustrating difference between pre-and post-intervention [32], and will also presented descriptive summary of the primary and secondary outcomes in tables.

## Data Availability

N/A

## Acknowledgements

We thank the funders.

## Notes

### Competing Interest Statement

The authors have declared no competing interest.

### Funding Statement

The ELEPHANT project is funded by the National Institute for Health and Care Research (NIHR) Public Health Research programme (NIHR129885; https://fundingawards.nihr.ac.uk/award/NIHR129885). The views expressed are those of the authors and not necessarily those of the NIHR or the Department of Health and Social Care.

